# Treatment Patterns and Outcomes of Pediatric Rhabdomyosarcoma in a Resource-limited setting:A Retrospective study from a Tertiary Hospital in Ethiopia

**DOI:** 10.64898/2026.10.29.26366799

**Authors:** Samson Zerihun, Sonia Worku Semayneh, Halid Worku Jemil, Birhanu Tesfaye Liyew, Adugna Fekadu, Damena Teshome Reta, Munir Awol Aman, Kidus Geabriel Yohannes, Gashaw Arega Mekonnen

## Abstract

**Background:** Rhabdomyosarcoma (RMS) is the most common pediatric soft tissue sarcoma. Despite excellent survival in high-income countries, data on treatment and outcomes remain limited in resource-limited settings. The aim of this study was to assess treatment patterns and survival outcomes of pediatric RMS in Ethiopia.

**Methods:** We conducted a retrospective study of newly diagnosed pediatric RMS patients evaluated at Tikur Anbessa Specialized Hospital from January 2021 to September 2025. Data were extracted from medical records, and treatment patterns and survival were assessed using descriptive and Kaplan–Meier analyses.

**Results:** A total of 94 pediatric patients with Rhabdomyosarcoma were included. The median age at diagnosis was 4.0 years (IQR: 3.0–6.3 years). Of these patients, 23.4% were classified as low-risk, 61.7% as intermediate-risk, and 14.9% as high-risk.Multimodality treatment was delivered to 59.6% of patients, while 33.0% received chemotherapy alone and 7.4% received no treatment. At diagnosis, curative treatment intent was planned for 83% of patients, but only 71.8% ultimately received it. Treatment received varied by risk group: most (90.0%) low-risk patients got curative treatment, while only 64.3% of intermediate-risk patients got curative treatment intent. No high-risk patients received curative treatment.At a median follow-up of 21.0 months (95% CI: 17.6–24.4 months), median overall survival and event-free survival were 23.0 and 15.0 months, respectively. The 1-year and 2-year overall survival rates were 70% and 42%, respectively, while the corresponding event-free survival rates were 59% and 33%. In multivariable Cox regression, treatment modality (p < 0.001) and total chemotherapy cycles (p =0.001) remained independently associated with overall survival, while risk group was not significant (p =0.14).

**Conclusion:** Pediatric rhabdomyosarcoma in Ethiopia presents with a high rate of high-risk disease. Multimodality therapy and completion of chemotherapy cycles are key determinants of improved survival. These findings highlight the urgent need for early detection, improved access to multimodal treatment, and strategies to reduce treatment abandonment and improve treatment completion rates in resource-limited settings. Larger prospective studies are warranted to validate these findings and guide evidence-based practice.

## Introduction

Rhabdomyosarcoma (RMS) is a malignant soft tissue sarcoma arising from primitive mesenchymal cells that are committed to skeletal muscle differentiation [1]. It is the most common soft tissue sarcoma in children, accounting for approximately 50% of all pediatric soft tissue sarcomas. However, RMS is a rare malignancy overall, representing only 3–4% of all childhood cancers [2, 3].

Historically, surgery alone cured fewer than 20% of patients with RMS, reflecting the high prevalence of occult micrometastatic disease. With the development of multimodal treatment protocols by international cooperative groups, outcomes have substantially improved, with more than 70% of children with localized RMS achieving long-term cure. Prognosis varies by risk group: low-risk patients have an expected event-free survival of 70–85% or greater, intermediate-risk patients 50–70%, and high-risk patients less than 30%. Current management incorporates chemotherapy for systemic disease control and cytoreduction, surgery when feasible, and radiotherapy for local control of residual or microscopic disease [4–10]. However, effective delivery of multimodal therapy requires coordinated multidisciplinary team (MDT) care and adequate healthcare resources, including access to chemotherapy, surgery, radiotherapy, pathology, and supportive care [11, 12].

Evidence on treatment patterns and associated outcomes of pediatric RMS in resource-limited settings, including sub-Saharan Africa and Ethiopia, where the delivery of multimodal therapy remains challenging, is limited and largely derived from small, single-center studies [13–16]. This study therefore aimed to assess treatment patterns and survival outcomes of pediatric RMS at a tertiary hospital in Ethiopia. The findings may inform treatment decision-making, resource allocation and provide a foundation for future multicenter prospective studies in the country and sub-Saharan Africa.

## Materials and Methods

### Study design and setting

This retrospective study was conducted on pediatric patients (<18 years of age) diagnosed with rhabdomyosarcoma (RMS) by treating physician and/or multidisciplinary team (MDT) based on histopathological and imaging findings between January 1, 2021, and September 30, 2025, who were planned for treatment at Tikur Anbessa Specialized Hospital. During this study period, Tikur Anbessa Specialized Hospital was one of the few centers in Ethiopia providing comprehensive cancer care, including radiotherapy (RT) services. The reporting of this study adheres to the Strengthening the Reporting of Observational Studies in Epidemiology (***STROBE***) guidelines[17].

### Study population, sample size and eligibility criteria

A census sampling approach was used; therefore, no formal sample size calculation was performed. All consecutive, newly diagnosed pediatric patients (<18 years of age) with rhabdomyosarcoma (RMS) who were evaluated and planned for treatment at Tikur Anbessa Specialized Hospital (TASH) between 1 January 2021 and 30 September 2025 were enrolled, regardless of whether the intended treatment was ultimately delivered. Diagnosis of RMS was established by the treating physician and/or multidisciplinary team (MDT) on the basis of compatible histopathological and image findings. The patient selection process and reasons for exclusion are summarized in (Fig 1).

**Fig 1.**
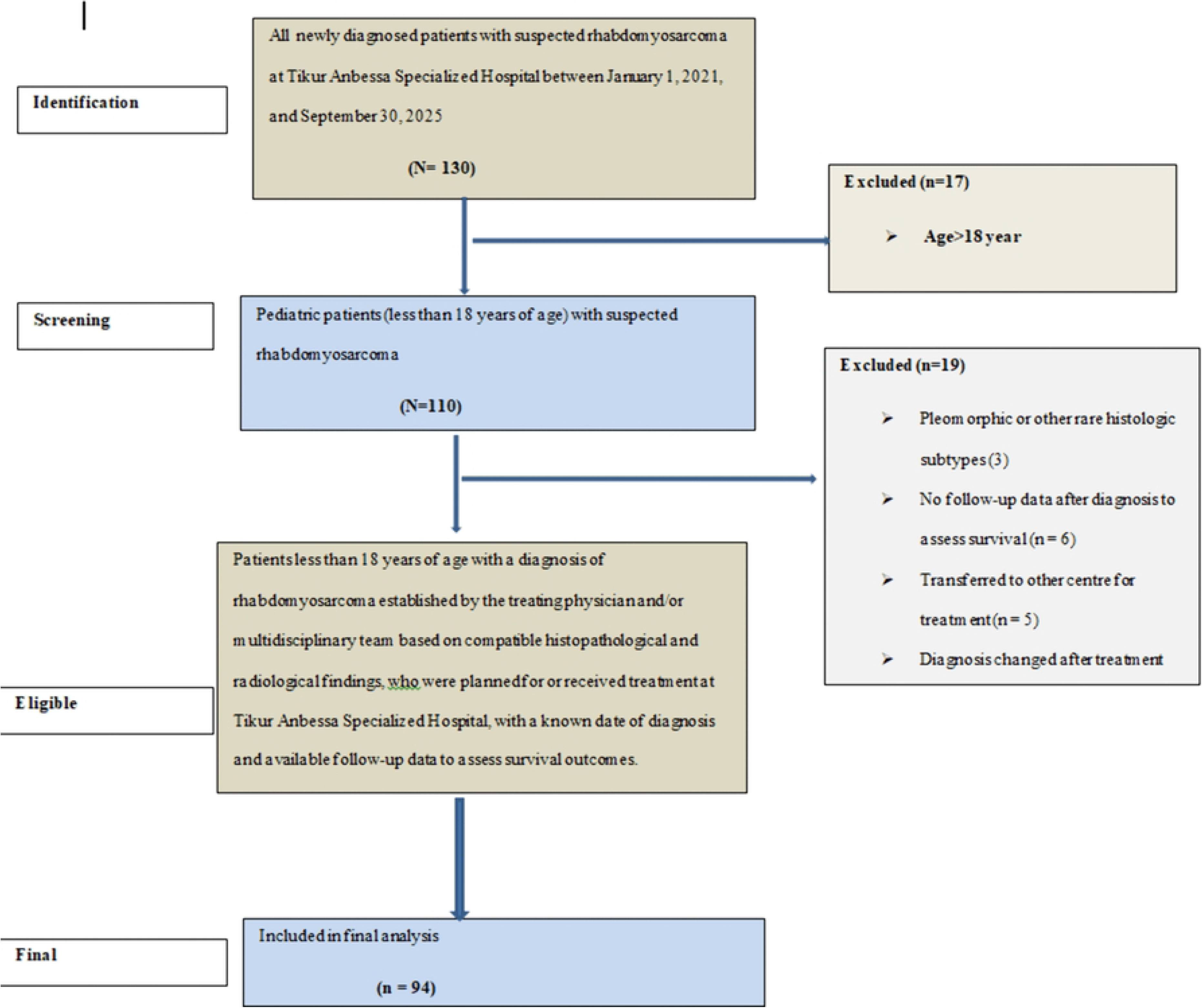
A flow diagram summarizing the patient selection process. Of 130 patients initially identified, 17 were excluded due to age greater than 18 years. Among the remaining 113 pediatric patients, 19 were excluded for various reasons (Pleomorphic or other rare histologic subtypes, no follow-up data, transfer to another centre, diagnosis changed, or unknown date of diagnosis), resulting in a final cohort of 94 patients included in the analysis.

### Diagnostic approach

As this study was conducted in a resource-limited setting with limited access to immunohistochemistry and molecular testing, the diagnosis of rhabdomyosarcoma (RMS) was established by the treating physician and/or multidisciplinary team (MDT) based on compatible histopathologic and radiologic findings. Molecular studies for diagnostic confirmation or risk stratification were not performed due to the unavailability of these tests in our setting. Similarly, immunohistochemistry (IHC) markers for RMS were not mandatory and were not part of standard practice during the study period. All cases included in the final analysis were diagnosed and managed as RMS by the treating physician and/or MDT based on the available clinical, histopathological, and radiological data. Given the absence of IHC and molecular confirmation, we included the following histologic subtypes in this study: embryonal RMS (including botryoid and spindle cell variants), alveolar RMS, and sclerosing RMS, as diagnosed by the treating physician and/or multidisciplinary team (MDT). Pleomorphic RMS and other rare histologic subtypes were excluded due to the diagnostic uncertainty that may arise without ancillary IHC or molecular studies.

### Data collection and quality assurance

The data collection time was between September 1,2025 and October 30,2025.The data were collected using KoboToolbox. Data collectors were trained before the start of the data collection and supervised routinely during data collection processes. Data were reviewed for completeness and clarity routinely by primary investigator.

### Treatment classification and definitions

For the purposes of this study, treatment intent was classified as curative or palliative based on the documented treatment plan and the intent of treatment actually delivered. Curative intent referred to treatment planned or administered with the goal of achieving definitive disease control or cure, whereas palliative intent referred to treatment planned or administered primarily to relieve symptoms or control disease without intent to achieve cure.

Treatment received was categorized as multimodality treatment when two or more treatment modalities (chemotherapy, surgery, and/or radiotherapy) were delivered and as single-modality treatment when only one modality was delivered. Patients who abandoned treatment before receiving any anticancer therapy were classified as having received no treatment.

Surgical treatment was classified as complete resection (tumor removal with negative margins), partial resection (gross residual disease with visible tumor left behind, or microscopic residual disease with positive margins), or biopsy only (diagnostic sample obtained without tumor removal).

Chemotherapy data included whether chemotherapy was administered and the total number of cycles received. Curative (radical) treatment was defined as oncologic treatment delivered with curative intent and aimed at complete tumor eradication. Palliative treatment was defined as non-curative oncologic treatment intended primarily for symptom control and/or prolongation of survival.

Radiotherapy indication was determined from the documented treatment plan and classified as planned with curative intent, planned with palliative intent, or not planned/not documented. Radiation dose was reported in Gray (Gy).

### Risk Group Classification

Clinical staging was performed according to the TNM staging system which incorporating the primary tumor size and anatomic site (T), regional lymph-node involvement (N), and presence or absence of distant metastases (M) [18]. Intergroup Rhabdomyosarcoma Study Group (IRSG) Clinical Group was based on the extent of surgical resection: Group I (completely resected), Group II (grossly resected with microscopic residual), Group III (incomplete resection or biopsy with gross residual), and Group IV (distant metastatic disease at presentation)[19]. In RMS risk group classification incorporates multiple established prognostic factors (histology type, clinical stage and clinical grouping) into a single composite variable that forms the basis for treatment stratification. Despite current risk stratification systems that incorporate molecular factors such as PAX3/7-FOXO1 fusion status [20], we used the pre-molecular era, clinical and histopathological based classification system because molecular testing was not available, not performed, or not documented at our centre during the study period [19].

### Response assessment and follow-up

Treatment response was assessed retrospectively using RECIST criteria (version 1.1) based on radiology reports. Radiologic assessment was performed using computed tomography (CT) scans or magnetic resonance imaging (MRI), depending on availability. Response categories were defined as: complete response (CR), partial response (PR), stable disease (SD), or progressive disease (PD) [21]. Response assessment was performed at 4 to 12 weeks after completion of the last treatment modality.

Follow-up evaluation, including imaging, was performed at variable intervals based on clinical indication and treating physician discretion in routine practice. Recurrence or disease progression was identified from clinical evaluation and radiological findings documented in patient records.

### Outcome Definitions

Overall Survival (OS) was defined as the time from date of diagnosis of rhabdomyosarcoma to death from any cause. Patients alive at last follow-up or lost to follow-up were censored at the date of last contact.

Event-free survival (EFS) was defined as the time from diagnosis to the first occurrence of any of the following events: disease progressive, disease recurrence, or death from any cause.

### Statistical Analysis

Descriptive statistics were used to summarize baseline demographic, clinical, and treatment characteristics. Continuous variables were summarized using mean or median, as appropriate, and categorical variables were presented as frequencies and proportions (%). Overall survival (OS) and event-free survival (EFS) were estimated using the Kaplan–Meier method, and survival distributions were compared using the log-rank test. Univariable and multivariable Cox proportional hazards regression was performed to identify factors independently associated with OS and EFS. Given the limited sample size and number of events (40 deaths for OS and 53 events for EFS), the multivariable models were restricted to three prespecified clinically relevant variables: treatment modality, risk group, and total chemotherapy cycles. This yielded events-per-variable (EPV) ratios of 13.3 for OS and 17.7 for EFS. The proportional hazards assumption was assessed using Schoenfeld residuals. The global test was not statistically significant (χ² = 0.95, p = 0.96). A two-sided p value < 0.05 was considered statistically significant. All analyses were performed using IBM SPSS Statistics version 27 and Stata version 15.

### Ethics approval and consent to participate

Ethical approval was obtained from the Departmental Research and Ethics Committee of the Department of Hematology, Oncology, and Nuclear Medicine, Addis Ababa University College of Health Sciences, before initiation of the study. The study was conducted in accordance with the principles of the 1964 Declaration of Helsinki and its later amendments. Due to the retrospective nature of the study and the use of anonymized patient records, the requirement for informed consent to participate was waived by the Departmental Research and Ethics Committee of the Department of Hematology, Oncology, and Nuclear Medicine, Addis Ababa University College of Health Sciences. The authors ensured that the rights of the patients were not violated in any manner.

## Results

A total of 113 pediatric patients with a diagnosis of rhabdomyosarcoma established by the treating physician and/or multidisciplinary team based on compatible histopathologic and radiologic findings, which were evaluated between January 2021 and September 2025, were reviewed using the electronic medical record system and patient charts. After applying the eligibility criteria, 94 patients were included in the final analysis (Fig 1).

### Patient and Disease Characteristics

The median age was 4.0 years (IQR, 3.0–6.3 years), with an age range of 0.3–17.0 years.The most common age groups of patients diagnosed with RMS were 1-5 years old.The most common primary tumor site was genitourinary (27.7%), followed by non-parameningeal head and neck site (22.3%). Regarding histologic type, embryonal subtype was the most frequent (n=60, 63.8%), followed by alveolar (n=30, 31.9%). The vast majority of patients (77.7%) diagnosed with large primary tumor size (≥5cm). Detailed baseline characteristics are shown in (**Table 1**).

**Table 1.** Demographics and Clinical Characteristics of RMS Patients (Age ≤ 18 Years).

| **Variables** | **Category** | **Number (n)** | **Proportion (%)** |
|---|---|---|---|
| **Age at diagnosis**<br>Range ( 0.3-17 years) | ≤1 year | 7 | 7.4 |
|  | 1-5 year | 50 | 53.2 |
|  | 5-10 year | 29 | 30.9 |
|  | ≥10 year | 8 | 8.5 |
| **Sex** | Male | 50 | 53.2 |
|  | Female | 44 | 46.8 |
| **Primary tumor anatomic site** | Orbit | 14 | 14.9 |
|  | Parameningeal | 12 | 12.8 |
|  | Non-parameningeal | 21 | 22.3 |
|  | Genitourinary | 26 | 27.7 |
|  | Extremity | 8 | 8.5 |
|  | Other | 13 | 13.8 |
| **Primary tumor site classification** | Favorable* | 47 | 50.0 |
|  | Unfavorable\$ | 47 | 50.0 |
| **Primary tumor size** | <5cm | 21 | 22.3 |
|  | ≥5cm | 73 | 77.7 |
| **Histologic subtype** | Embryonal | 60 | 63.8 |
|  | Alveolar | 30 | 31.9 |
|  | Spindle cell | 3 | 3.2 |
|  | Botryoid | 1 | 1.1 |

\*Favorable primary tumor sites include orbit, non-parameningeal head and neck, and genitourinary (non-bladder/prostate).
\$Unfavorable primary tumor sites include parameningeal, extremity, and other sites.

### Staging and Risk grouping results

Majority of Patients were diagnosed at advanced clinical stage and group, IRS stage III (58.5%),and Group III(55.9%).Risk grouping revealed that intermediate risk group were more common (61.7%),followed by low risk (23.4%),and rarely high risk(14.9%).(**Table 2**)

**Table 2.** Pretreatment clinical staging and Risk grouping of RMS patients,Age ≤ 18 Years.

| **Variables** | **Category** | **Number (n)** | **Proportion (%)** |
|---|---|---|---|
| **IRS<sup>a</sup> stage at diagnosis** | I | 13 | 13.8 |
|  | II | 12 | 12.8 |
|  | III | 55 | 58.5 |
|  | IV | 14 | 14.9 |
| **Clinical Group** | Group I | 11 | 11.8 |
|  | Group II | 17 | 18.3 |
|  | Group III | 52 | 55.9 |
|  | Group IV | 13 | 14.0 |
|  | unknown | 1 | 1.1 |
| **Risk Group<sup>b</sup>** | Low risk | 22 | 23.4 |
|  | Intermediate risk | 58 | 61.7 |
|  | High risk | 14 | 14.9 |

<sup>a</sup>IRS, Intergroup Rhabdomyosarcoma Study.
<sup>b</sup>Risk group classification was based on the pre-molecular era classification system, which includes clinical stage, clinical group, and histologic subtype.

### Treatment modalities

Of the 94 patients, 56 (59.6%) received multimodality treatment, 31 (33.0%) received single-modality treatment (all of which were chemotherapy alone), and 7 (7.4%) received no treatment (treatment abandonment). At diagnosis, 78 patients (83.0%) were initially planned for curative-intent treatment, whereas 17.0% were planned for palliative treatment. Among the 78 patients initially planned for curative intent, 71.8% were ultimately received curative treatment, 23.1% received palliative treatment, and 5.1% received no treatment. Treatment intent planned and delivered differed by risk group. All low-risk patients were initially planned for curative treatment intent, with 90.9% ultimately receiving curative treatment. Among intermediate-risk patients, 96.6% were initially planned for curative treatment intent; of these, 64.3% received curative treatment, 30.4% received palliative treatment, and 5.4% received no treatment. In contrast, all high-risk patients were initially planned for palliative and none received curative treatment.

Treatment modality received varied by risk group (Fisher-Freeman-Halton exact test, p < 0.001) (Table 3). Among the 56 patients who received multimodality treatment, most were intermediate-risk (64.3%), followed by low-risk (35.7%); no high-risk patients received multimodality treatment. Regarding the specific multimodality regimens, low-risk patients most commonly received chemotherapy plus surgery (50.0%), whereas intermediate-risk patients most commonly received chemotherapy plus radiotherapy (55.6%).

**Table 3:** Treatment Patterns by Risk Group.

| **Variables** | **Low Risk (n=22)** |  | **Intermediate Risk (n=58)** |  | **High Risk (n=14)** |  |
|---|---|---|---|---|---|---|
|  | **n** | **%** | **n** | **%** | **n** | **%** |
| **Multimodality** | **20** | **35.7** | **36** | **64.3** | **0** | **(0.0)** |
| Chemotherapy + radiotherapy | 2 | 9.1 | 20 | 90.9 | – | – |
| Chemotherapy + Surgery | 10 | 50.0 | 10 | 50.0 | – | – |
| Chemotherapy radiotherapy +Surgery | 8 | 57.1 | 6 | 42.9 | – | – |
| **Single modality** | **1** | **3.2** | **18** | **58.1** | **12** | **(38.7)** |
| **No treatment** | **1** | **14.3** | **4** | **57.1** | **2** | **28.6** |

### Surgical treatment

Out of 94 patients, 29 (30.9%) underwent complete resection, 5 (5.3%) underwent partial resection, and 60 (63.8%) underwent biopsy only.

### Chemotherapy

Chemotherapy was administered to 87 patients (92.6% of the total cohort). The median number of chemotherapy cycles received was 36 (IQR: 18–39), with a minimum of 4 cycles and a maximum of 42 cycles. The number of cycles differed by treatment modality. Among the 56 patients who received multimodality treatment, the median number of chemotherapy cycles was 39 (range: 12–42).In contrast, among the 31 patients who received single-modality treatment, the median number of cycles was significantly lower at 16 (range: 4–39).

### Radiotherapy

Radiotherapy was planned for 90 patients (95.7%), while it was not planned or not documented in 4.3%. Among the 90 patients with a radiotherapy plan, 83.3% had curative treatment intent and 16.7%) had palliative treatment intent. Overall, radiotherapy was administered to 42 patients (44.7% of the total cohort). Among the 42 patients who received radiotherapy, 85.7% of them received it with curative intent. Among the 75 patients with curative/radical intent plan, 48.0% received radical radiotherapy.

Among the 42 patients who received radiotherapy, the median time from surgery or biopsy to radiotherapy was 35.5 weeks (4-93 week). For the 36 patients treated with curative intent, the median time was 41.0 weeks (21-93 week).Among the 36 patients who received curative intent radiotherapy, the median total dose was 47.7Gy with the range of 36.0–50.4Gy.

### Treatment response

Treatment response was assessed in 77 patients (81.9%) using contrast-enhanced computed tomography (CECT) or magnetic resonance imaging (MRI). The response assessment was performed at 4 to 12 weeks after completion of treatment. Among the evaluated 77 patients, 40.3% achieved complete response, 22.1% had partial response, 13.0% had stable disease, and 24.7% had progressive disease. The overall response rate (completes response + had partial response) was 62.4%

Among the 56 patients who received multimodality treatment, 52 (92.9%) were evaluated for response. Of these, evaluated for response, 59.6% achieved complete response, 26.9% had partial response, 5.8% had stable disease, and 7.7% had progressive disease. Among the 31 patients who received single-modality treatment, 25 (80.6%) were evaluated for response. Of these evaluated for response, 12.0% achieved PR, 28.0% had SD, and 60.0% had PD.

### Overall survival

At the time of last follow-up or mortality check, 54 patients (57.4%) were censored (34 alive, 20 lost to follow-up), while 40 patients (42.6%) had died. The median follow-up time for the entire cohort, estimated using reverse Kaplan-Meier analysis, was 21.0 months (95% CI: 17.6–24.4 months). Follow-up duration ranged from 1 to 52 months.

The median overall survival for the entire cohort was 23.0 months (95% CI: 7.5–38.5 months). The Kaplan-Meier estimated 1 and 2-year overall survival rates, were 70% and 42%, respectively (**Fig 2**).

**Fig 2.**
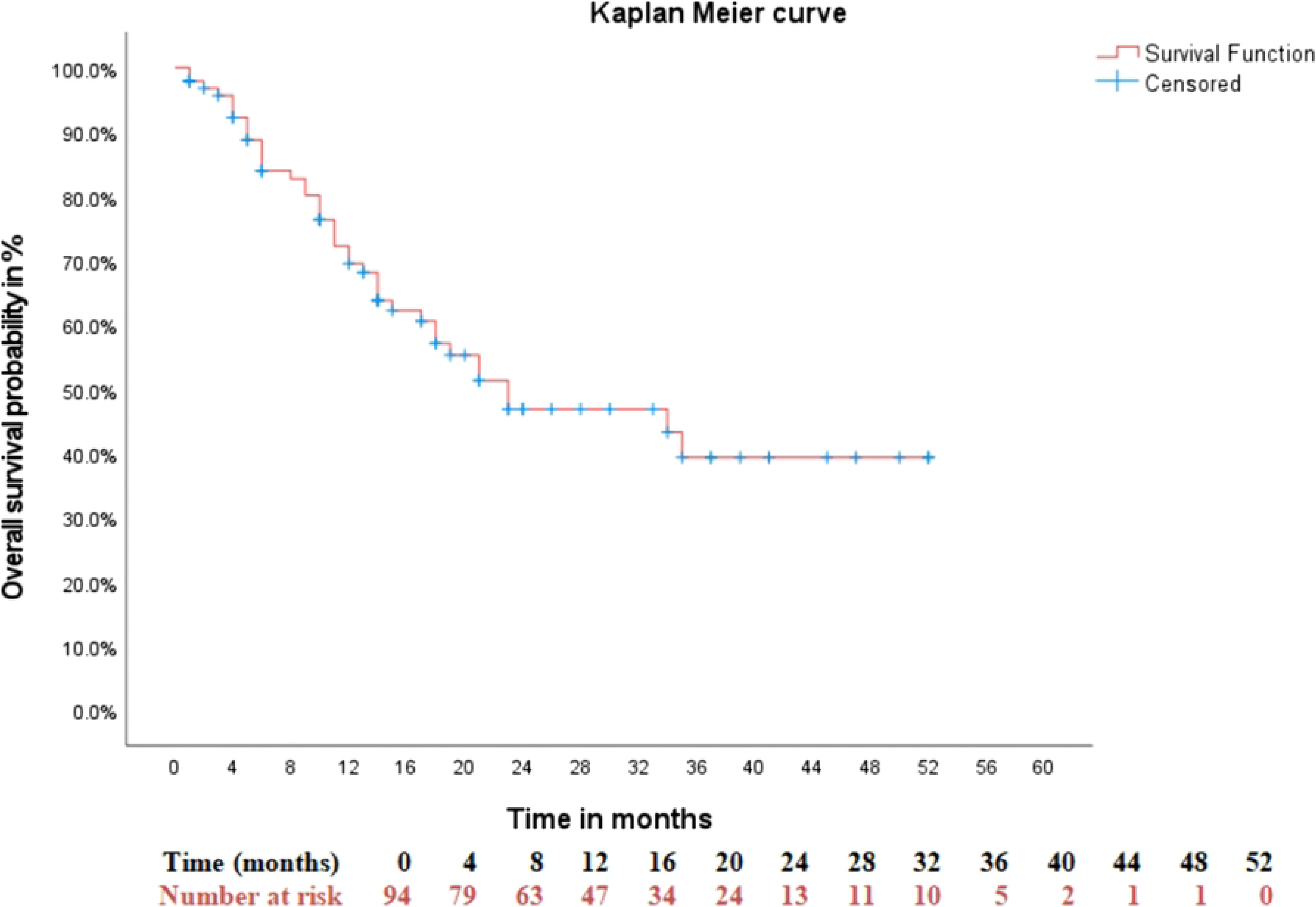
Kaplan–Meier overall survival curve for the entire cohort of patients with rhabdomyosarcoma. The median overall survival was 23.0 months (95% CI: 7.5–38.5 months). The Kaplan Meier estimated 1-year and 2-year overall survival rates were 70% and 42%, respectively.

Kaplan-Meier analysis demonstrated a significant difference in overall survival according to treatment modality (log-rank, p < 0.001). The median overall survival was not reached for multimodality patients, compared with 10.0 months (95% CI: 6.7–13.3) for single-modality patients and 3.0 months for those receiving no treatment. The 1-year and 2-year OS rates were 88% and 66% for multimodality, 37% and 0% for single-modality, and 0% and 0% for no treatment (**Fig 3**).

**Fig 3.**
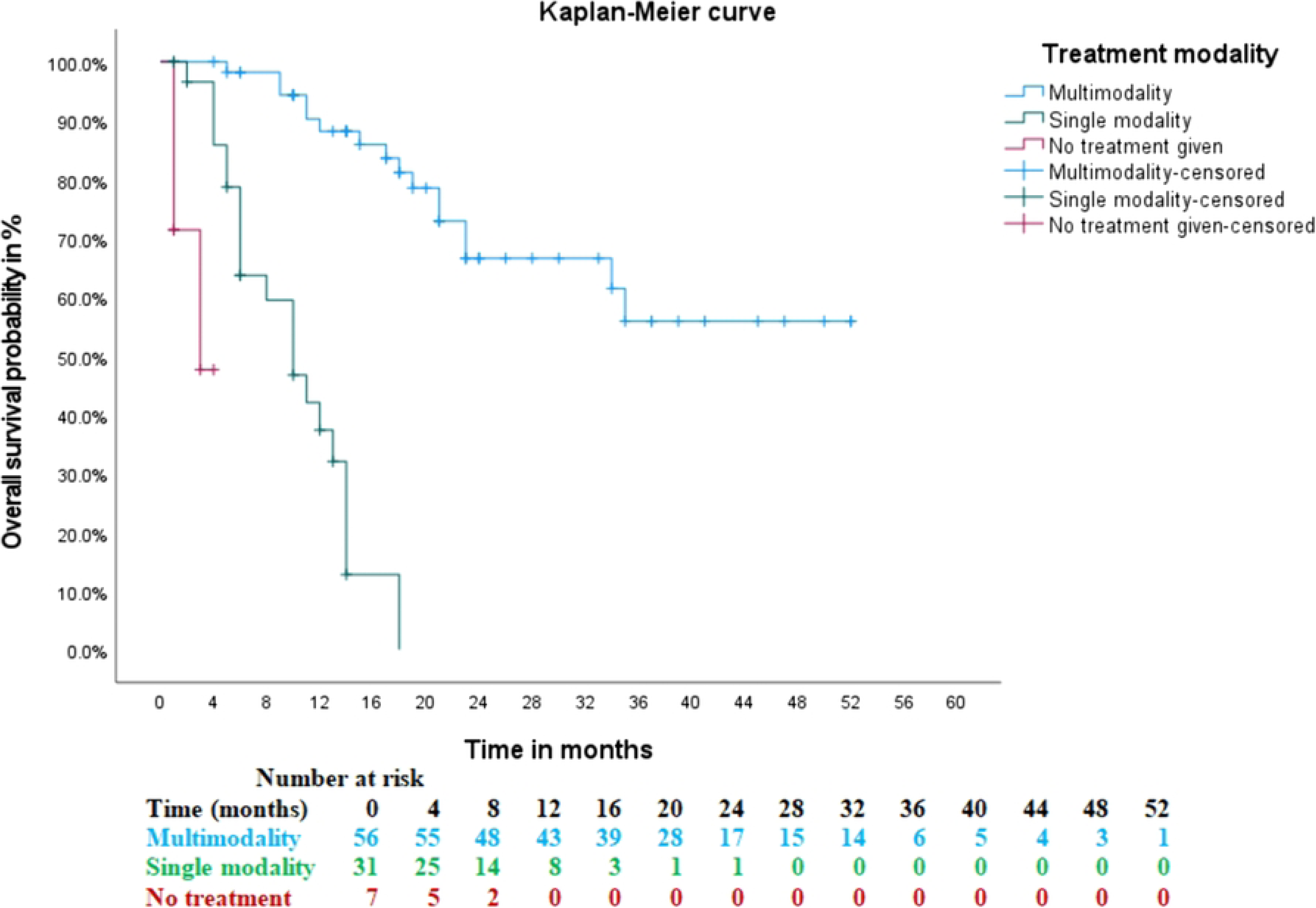
Kaplan–Meier overall survival stratified by treatment modality. Overall survival differed significantly across treatment groups (log-rank p < 0.001). The 1-year and 2-year OS rates were 88% and 66% for multimodality, 37% and 0% for single-modality, and 0% and 0% for no treatment

Overall survival differed significantly across risk groups (log-rank test, p < 0.001). The median overall survival was not reached in the low-risk group, 23.0 months (95% CI: 17.7–28.3 months) in the intermediate-risk group, and 5.0 months (95% CI: 0.7–9.3 months) in the high-risk group. The 1-year and 2-year overall survival rates for low-risk patients were 90% and 78%, respectively; for intermediate-risk patients, 77% and 42%; and for high-risk patients, 18% and 0%.

### Event Free Survival (EFS)

For the entire cohort, the median event-free survival was 15.0 months (95% CI: 9.6–20.4 months). The 1-year and 2-year event-free survival rates, estimated using the Kaplan-Meier method, were 59% and 33%, respectively (**Fig 4**)

**Fig 4:**
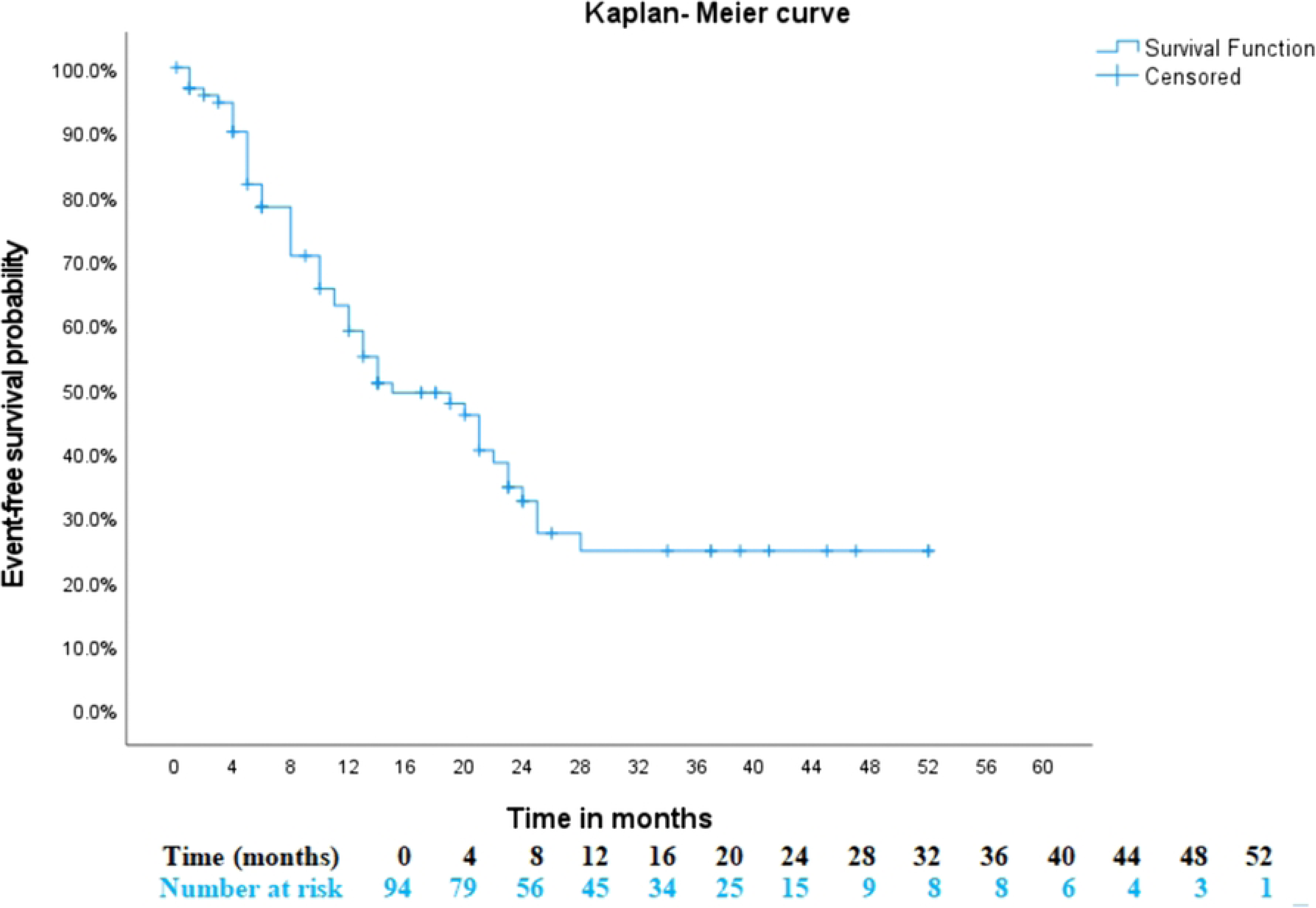
Overall event-free survival in pediatric patients with rhabdomyosarcoma. The median event-free survival was 15.0 months (95% CI: 9.6–20.4 months), with 1-year and 2-year survival rates of 59% and 33%, respectively.

Event-free survival differed significantly across risk groups (log-rank test, p < 0.001). The median event-free survival for low-risk patients was 25.0 months (95% CI: 20.3–29.7 months), 15.0 months (95% CI: 9.6–20.4 months) for intermediate-risk patients, and 5.0 months (95% CI: 3.8–6.2 months) for high-risk patients. The 1-year and 2-year event-free survival rates for low-risk patients were 90% and 78%, respectively; for intermediate-risk patients, 60% and 24%; and for high-risk patients, 5% and 0%

Event-free survival differed significantly by treatment modality (log-rank test, p < 0.001). The median EFS was 24.0 months (95% CI: 21.2–26.8) for multimodality patients, 6.0 months (95% CI: 3.6–8.4) for single-modality patients, and 4.0 months (95% CI: 0.9–7.1) for those receiving no treatment. The 1-year and 2-year EFS rates were 85% and 50% for multimodality, 16% and 0% for single-modality, and 0% and 0% for no treatment (Fig 5).

**Fig 5.**
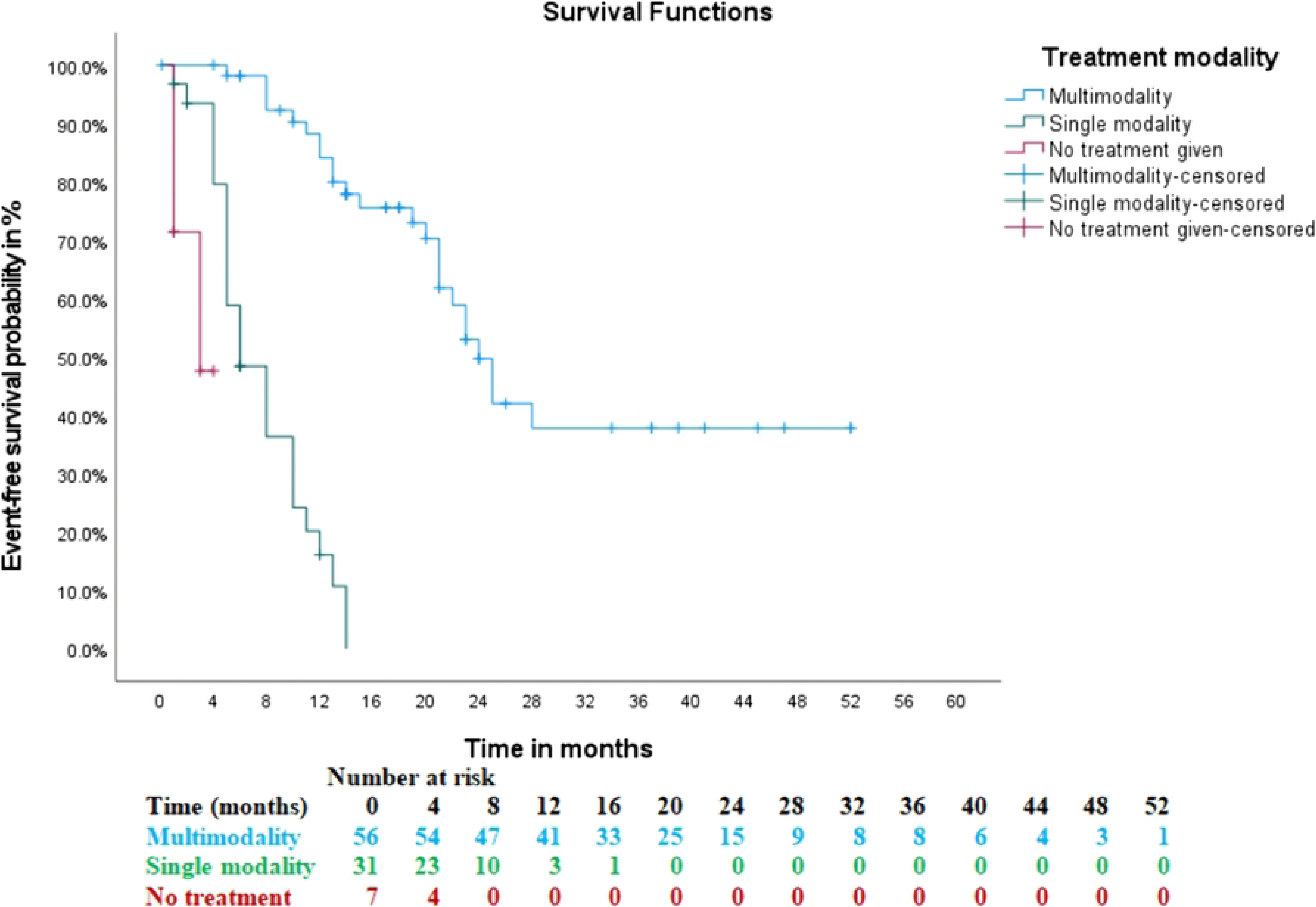
Kaplan–Meier event-free survival stratified by treatment modality. The 1-year and 2-year EFS rates were 85% and 50% for multimodality, 16% and 0% for single-modality, and 0% and 0% for no treatment.

Given that 20 patients (21.3%) were lost to follow-up, a sensitivity analysis was performed to assess the robustness of the survival estimates, reclassifying these patients as having died at the time of their last contact. The median overall survival (OS) was 17.0 months (95% CI: 12.1–21.9 months) in the sensitivity analysis. Although the point estimate shifted downward (17 months from 23 months), the confidence intervals overlap substantially, indicating that the difference is not statistically significant. This suggests that the survival estimates are relatively robust and that the loss to follow-up did not materially bias the overall findings.

### Prognostic factors

To identify prognostic factors for overall survival, we performed univariable and multivariable Cox regression analyses. In univariable analysis, treatment modality, risk group, and total chemotherapy cycles were each significantly associated with survival (p < 0.05, for all). In multivariable analysis, treatment modality (HR = 5.02; 95% CI: 1.79–14.06; p = 0.002) and total chemotherapy cycles (HR = 0.95; 95% CI: 0.91–0.98; p = 0.001) remained independently associated with overall survival. The intermediate-risk group showed a non-significant trend toward higher mortality (HR = 2.14; 95% CI: 0.77–5.95; p = 0.14), while the high-risk group had a significantly elevated risk (HR = 5.35; 95% CI: 1.47–19.55; p = 0.011), though the wide confidence intervals for both estimates indicate reduced precision and warrant cautious interpretation **(S1 Table).**

To identify prognostic factors for event-free survival (EFS), we performed univariable and multivariable Cox regression analyses. In univariable analysis, treatment modality, risk group, and total chemotherapy cycles were each significantly associated with EFS (all p < 0.05). In multivariable analysis, treatment modality and total chemotherapy cycles remained independently associated with EFS, while risk group was not statistically significant. Single-modality treatment was associated with a 6-fold higher risk of an event compared with multimodality treatment (HR = 6.09; 95% CI: 2.49–14.90; p < 0.001), and each additional chemotherapy cycle reduced event risk by 6% (HR = 0.94; 95% CI: 0.92–0.97; p < 0.001). Although risk group was significant in univariable analysis, it did not retain statistical significance in multivariable analysis for intermediate-risk (HR = 1.66; 95% CI: 0.73–3.76) and high-risk (HR = 2.41; 95% CI: 0.80–7.29). The wide confidence intervals suggest that the study was likely underpowered to detect a true independent effect **(S2 Table).**

## Discussion

To the best of our knowledge, this is the first study from Ethiopia to comprehensively describe both treatment patterns and survival outcomes in pediatric rhabdomyosarcoma (RMS). While a previous study from our center described the clinicopathological characteristics of 50 patients, survival data have not been reported from Ethiopia. Regionally or from sub-Sahara Africa, only a few small studies from Uganda, Rwanda and Nigeria have reported survival outcomes, with poor 5-year survival rates and high treatment abandonment [13–16]. This study addresses this critical evidence gap by providing essential baseline data on treatment approaches and survival outcomes in a resource-limited setting. These findings establish a benchmark for comparison with international data and lay the foundation for future prospective multicenter studies in the country and region.

In this study, the median age at diagnosis was 4 years, and males comprised 53.2% of the cohort. These findings are consistent with previous reports from Ethiopia (median: 3 years; male: 54%) and other Sub-Saharan African studies. Advanced-stage disease at presentation was predominant, with Stage III disease in 58.5% of patients and Clinical Group III in 55.9%. This distribution is consistent with reports from other Sub-Saharan African studies, such as a Ugandan study in which 90.6% of patients presented as Group III, reflecting the common challenge of delayed presentation across the region [14, 16, 22].

The current standard of care for RMS is complex and risk-adapted, involving multimodal treatment with chemotherapy, surgery when feasible, and radiotherapy, guided by multidisciplinary assessment. These recommendations are based primarily on the results of the Intergroup Rhabdomyosarcoma Study Group (IRSG) protocols, including IRS-IV, IRS-V (D series), and the Children’s Oncology Group (COG) ARST (Adolescent and Young Adult Rhabdomyosarcoma and Soft Tissue Sarcoma Trial) protocols [12, 23–25]. Treatment intensity and modality are tailored according to established clinicopathologic risk factors. Low-risk patients generally receive less intensive chemotherapy, whereas intermediate- and selected high-risk patients receive more intensive regimens. Surgery is central to the management of local control, with complete excision recommended when feasible; for unresectable tumors, biopsy is followed by neoadjuvant chemotherapy and local therapy. Radiotherapy is indicated for all patients except those with embryonal/fusion-negative Clinical Group I tumors, with doses of 41.4–45 Gy for microscopic residual disease and 45–50.4 Gy for gross residual disease, typically administered after four cycles of chemotherapy[11, 12, 23–25]. Although the detailed treatment algorithm is complex and beyond the scope of this summary, these principles represent the current standard multimodality approach to RMS.

In our cohort, although chemotherapy was initiated in 92.6% of cases, only 59.6% received multimodality treatment, and radiotherapy, planned for 95.7%, was delivered to only for 44.7%, highlighting significant gaps in local therapy delivery. Treatment deviations were notable by risk group, with curative rates dropping from 90.9% in low-risk to 64.3% in intermediate-risk patients, while all high-risk patients received palliative or no care. Prolonged radiotherapy waiting times and financial constraints are likely contributing factors. For 36 patients treated with curative intent, the median time was 41.0 weeks (21-93 week). This far exceed the recommended 6–14 week window for definitive local therapy in pediatric rhabdomyosarcoma. Compared to prior Ethiopian and Ugandan eport, radiotherapy delivery (53.2%) and multimodality rates have improved, and treatment abandonment (7.4%) was far lower than the 40% reported in Nigeria[14, 16, 22]. Overall, these findings suggest that while initiation of systemic therapy is relatively high, timely integration of definitive local therapy remains a major challenge in our setting.

In our cohort, the median overall survival (OS) was 23.0 months, with 1-year and 2-year OS rates of 70% and 42%. Median OS was not reached for patients receiving multimodality treatment, compared to 10.0 months for single-modality chemotherapy and 3.0 months for no treatment (p < 0.001). By risk group, median OS was not reached for low-risk, 23.0 months for intermediate-risk, and 5.0 months for high-risk patients (p < 0.001). The median event-free survival (EFS) was 15.0 months, with 1-year and 2-year EFS rates of 59% and 33%. Median EFS was 24.0 months for multimodality, 6.0 months for single-modality, and 4.0 months for no treatment (p < 0.001). By risk group, median EFS were 25.0 months for low-risk, 15.0 months for intermediate-risk, and 5.0 months for high-risk (p < 0.001). A sensitivity analysis excluding the 7 untreated patients (n = 87) showed 1-year and 2-year OS rates of 71% and 48%.

Our survival outcomes, while lower than those reported in Western cooperative group studies, are generally favorable relative to several reports from resource-limited settings. Compared with Uganda, where 1-year and 5-year OS were 65% and 12%, our cohort demonstrated higher short-term survival, although longer-term comparison is limited by differences in follow-up duration. Our 2-year OS of 42% was lower than the 48% reported from India, while our 2-year EFS of 33% exceeded the 22% reported in that study. The Nigerian study reported a median survival of 45 weeks, considerably shorter than the 23-month median OS observed in our cohort. In contrast, outcomes remain below data from major cooperative group trials that set benchmark survival estimates. For the low-risk group, the ARST0331 study reported 1- and 2-year EFS rates of 100% and 99% (Walterhouse et al., 2014). For the intermediate-risk group, the D9803 study reported 1- and 2-year EFS rates of 90% and 75% (Wolden et al., 2015). For the high-risk group, the IRS-IV study reported 1- and 2-year EFS rates of 62% and 45% (Breneman et al., 2003) [14, 22–26].

However, survival estimates in our study are limited by few patients at risk at 2 years, with 2-year survival reaching 0% in single-modality, no-treatment, and high-risk groups. Wide confidence intervals for median OS and EFS in high-risk subgroups also reflect small sample sizes and reduced precision. These findings warrant cautious interpretation and highlight the need for larger studies.

In this study, univariable analysis showed that treatment modality, risk group, and total chemotherapy cycles were each significantly associated with overall survival (OS) and event-free survival (EFS). In multivariable analysis, only treatment modality and chemotherapy cycles remained independent predictors. Compared with multimodality treatment, single-modality treatment was associated with a five-fold higher risk of death (HR = 5.02; 95% CI: 1.79–14.06; p = 0.002) and a six-fold higher risk of an event (HR = 6.09; 95% CI: 2.49–14.90; p< 0.001).

Estimates for the no-treatment group were extremely imprecise (OS: HR = 100.47, 95% CI: 7.37–1370.45; EFS: HR = 36.49, 95% CI: 4.57–291.36), reflecting the very small number of untreated patients (n = 7). Each additional chemotherapy cycle reduced the risk of death by 5% and the risk of an event by 6%. Risk group lost statistical significance after adjustment.

Interpretation of these findings is limited by the small sample size, particularly in the no-treatment and high-risk subgroups, resulting in wide confidence intervals and reduced statistical precision. The study may have been underpowered to detect an independent effect of risk group; therefore, the lack of statistical significance should be interpreted with caution, and the clinical importance of risk stratification cannot be excluded. While the point estimates suggest that treatment delivery and intensity may have been more immediate determinants of outcome than baseline risk in this setting, this observation remains exploratory and warrants confirmation in larger, prospective studies.

These results are consistent with evidence from Uganda and SEER analyses highlighting the survival benefit of combined local-control treatments. They suggest that, in this resource-limited setting, actual treatment delivery and intensity may have been more immediate determinants of outcome than baseline risk. Overall, the findings emphasize the critical importance of achieving timely, complete multimodality treatment in pediatric rhabdomyosarcoma, while underscoring the need for larger prospective studies to confirm these associations and better clarify the independent contribution of risk group [22, 26, 27].

## Conclusion

Overall, this study demonstrates that pediatric RMS in our setting is characterized by substantial treatment-delivery gaps, particularly in the timely provision of local therapy, despite relatively high chemotherapy utilization. Survival was better among patients receiving multimodality treatment and those completing more chemotherapy cycles, but remained below outcomes reported by international cooperative groups. These findings highlight the need to strengthen multidisciplinary, risk-adapted care and timely delivery of complete multimodality treatment to improve outcomes in resource-limited settings.

## Limitations of the study

This study has several limitations. First, its single-center retrospective design is subject to selection bias, information bias, and limitations inherent to the completeness and accuracy of routinely collected medical records. Second, the relatively small sample size (n=94) and limited number of observed events restricted the statistical power of survival analyses and limited the ability to perform comprehensive multivariable prognostic analyses involving multiple potential prognostic factors. Consequently, the multivariable findings should be interpreted cautiously and primarily as associations rather than causal relationships. Third, the relatively short follow-up period limited assessment of long-term survival and late treatment-related outcomes. Fourth, detailed information on chemotherapy regimens, dose intensity, treatment modifications, dose reductions, interruptions, and treatment-related toxicity was not sufficiently available for comprehensive analysis of treatment intensity and protocol adherence. Fifth, because of treatment allocation was not randomized and was influenced by disease characteristics, resectability, treatment availability, and patient-related factors, the observed association between treatment modality and survival may be affected by other unmeasured confounders. Finally, as this was a single-center study from a tertiary oncology institution, the findings may not be fully generalizable to other hospitals or regions in Ethiopia and sub-Saharan Africa. Larger multicenter prospective studies with longer follow-up, standardized treatment documentation, and more complete assessment of established prognostic factors are needed to validate these findings and better characterize determinants of survival in pediatric RMS in resource-limited settings.

## Author contributions

Ø **Conception/design**: Samson Zerihun, Halid Worku Jemil, and Sonia Worku Semayneh,Gashaw Arega Mekonnen

Ø **Literature search**- Samson Zerihun and Sonia Worku Semayneh and Birhanu Liyew,Damena Teshome Reta

Ø **Collection and/or assembly of data**: Samson Zerihun and Sonia Worku Semayneh, Birhanu Liyew

Ø **Data analysis and interpretation**: Samson Zerihun,Halid Worku Jemil, Adugna Fekadu, Sonia Worku Semayneh, Birhanu Liyew, Munir Aman, Kidus Yohannes,Gashaw Arega Mekonnen

Ø **Manuscript writing**: Halid Worku Jemil and Sonia Worku Semayneh, Birhanu Liyew, Kidus Yohannes,

Ø **Manuscript editing:** Samson Zerihun,Halid Worku Jemil, Adugna Fekadu, Sonia Worku Semayneh, Birhanu Liyew, Munir Aman, Kidus Yohannes,Gashaw Arega Mekonnen,Damena Teshome Reta

Ø **Final approval of manuscript**: All authors

## Data Availability

The minimal data set (and accompanying code, where generated) is available at (repository name) via (DOI hyperlink)

## Acknowledgements

The authors would like to acknowledge the staff and faculty of the Department of Hematology, Oncology, and Nuclear Medicine at Addis Ababa University, College of Health Sciences, for their continuous support during data collection and manuscript preparation. The authors also acknowledge the use of artificial intelligence–assisted language editing tools to improve the clarity and grammar of the manuscript. All scientific content, interpretations, and conclusions remain the responsibility of the authors.

## Funding

The author(s) have declared that no financial support was received for this research.

## Conflicts of interest

The authors have declared that no conflicts of interests exist.

## Consent for publication

Not applicable

## Abbreviations

RMS: Rhabdomyosarcoma
MDT: Multi-Displinary Team
RT: Radiation Therapy
TASH: Tikur Anbessa Specialized Hospital
IHC: Immunohistochemistry
Gy: Gray
TNM: Tumor-Node-Metastasis
IRSG: International Rhabdomyosarcoma Study Group
CT: Computerzed themography
MRI: Magnetic resonant image
CR: complete response
PR: partial response
SD: stable disease
PD: progressive disease
OS: overall survival
EFS: event free survival

## Supporting information

**S1Table.** Univariable and Multivariable Cox Regression Analysis for Prognostic Factors of Overall Survival of pediatrics rhabdomysarcoma.

**S2 Table.** Univariable and Multivariable Cox Regression Analysis of Prognostic Factors for Event Free Survival(EFS).

**S1 Data.**

## Notes

### Competing Interest Statement

The authors have declared no competing interest.

### Author Declarations

Treatment Patterns and Outcomes of Pediatric Rhabdomyosarcoma in a Resource-limited setting: A Retrospective study from a Tertiary Hospital in Ethiopia With Protocol Number HONM/11/18, was reviewed and approved by the Departmental Research Ethics Committee of the College of Health Sciences, Addis Ababa University. The protocol was discussed in Meeting No. HONM/11/18 and received ethical clearance on jun Given the retrospective nature of the study and use of anonymized patient records, the Committee approved a waiver of informed consent.

